# Obesity is associated with increased clearance and reduced drug exposure to metformin in youth with type 2 diabetes

**DOI:** 10.1101/2025.03.03.25323158

**Authors:** Shylaja Srinivasan, Alexander Floren, Kevin Yen, Eunsol Yang, Rada Savic

## Abstract

**Objective:** Youth with type 2 diabetes have higher metformin treatment failure rates when compared with adults. Our objectives were to develop a population pharmacokinetic (PK) model for metformin in youth to examine current dosing strategies and to quantify the relationship between metformin PK and obesity in youth with type 2 diabetes.

**Research Design and Methods:** We performed semi-intensive PK sampling in 50 youth less than 18 years with T2D on metformin. We measured metformin levels at pre-dose and at 1–2 and 3–6-hours after a supervised 1000 mg dose of immediate-release metformin. We used non-linear mixed effects modeling to develop a population PK model. Simulations were performed to estimate area under the 24-hour metformin concentration-time curve (AUC) values.

**Results:** Mean age was 15.0 (standard deviation (SD) 1.6) years, 66% were male, mean body mass index (BMI), 35.9 (SD 6.8) kg/m^2^, and mean estimated glomerular filtration rate (eGFR), 114.7 (SD 22.6) mL/min/1.73m^2^. Estimated clearance increased by 3% (21.4% relative standard error (RSE)) for every 1 kg/m^2^ increase in body mass index (BMI), and by 6% (53.8% RSE) for every 10 mL/min/1.73m^2^ increase in eGFR. When comparing AUC estimates from our model with data from 236 adults on metformin, median AUC for youth was lower than for adults on 2,000 mg of metformin (18.47 mg*h/L vs. 27.49 mg*h/L). Simulations showed that in youth, higher doses of at least 2,550 mg and up to 3,000 mg daily were needed to achieve exposures similar to adults.

**Conclusions:** Obesity is associated with increased metformin clearance and reduced AUC in youth and higher doses are needed to achieve comparable drug exposure to adults.

## Introduction

Type 2 diabetes once considered a disease of adulthood, is becoming increasingly frequent in youth with alarming predictions of future trends.^1-4^ Youth with type 2 diabetes have increased mortality and a very high incidence of microvascular complications affecting the eye, kidneys and nerves that can occur as early as ten years after diagnosis.^5^ It has therefore become critical that we employ effective treatment strategies to curb the progression of the disease right from the disease onset. Metformin remains first-line and is the foundation of treatment for type 2 diabetes in youth because it is safe, orally administered, low-cost, globally available, and well-tolerated. In addition, there is evidence from adult studies to suggest that there are benefits to metformin therapy beyond glycemic control including protection against diabetes related long-term cardiovascular, renal, and neurological complications. While there are newer medications such as the glucagon like peptide receptor agonist therapies which are now approved for use in youth with type 2 diabetes, they currently remain largely inaccessible due to formidable cost and supply-chain issues. Additionally, the long-term safety effects of these newer medications on youth remain largely unknown.

Despite metformin remaining the cornerstone of type 2 diabetes management, not all youth have sustained glycemic response to metformin and youth have worse glycemic response to metformin compared with adults. Metformin was first approved by the U.S. Food and Drug Administration (FDA) for pediatric use in children 10 years and older with type 2 diabetes in the year 2000 with dosing recommendations largely based on studies in adults and a small study in children. Currently, the maximum approved dose for metformin in children is 2,000 mg a day compared to a maximum approved dose of 2,550 mg in adults. While for most disease states, it is reasonable for pediatric dosing to be lower than in adults, this paradigm may not apply to type 2 diabetes. Type 2 diabetes in children is most often diagnosed in older children and adolescents after the onset of puberty and obesity may play a larger role in pediatric type 2 diabetes compared to the adult-onset form of the disease. For instance, the average body mass index (BMI) of youth participants with type 2 diabetes in the TODAY study at baseline was 34.9 kg/m^2^ and in the Restoring Insulin Secretion (RISE) study which evaluated the effect of medications on preserving β-cell function, the average BMI for youth participants at baseline was higher than that of the adults.

Pharmacokinetics (PK) quantifies the mathematical relationship between a drug’s administered dose and its concentration in the body, playing a crucial role in determining drug response. Understanding the sources of variability in metformin PK in youth with type 2 diabetes can inform simple dosing changes that can make this currently approved drug more effective. However, traditional ways of measuring PK are time consuming, requiring multiple blood draws and fraught with logistical constraints in children. Population PK modeling allows for evaluating population PK parameters and sources of variability in PK in a population of interest based on sparse and unbalanced PK data. Our objectives in this study were to evaluate the factors affecting the variability in metformin PK in youth with type 2 diabetes through the development of a population PK model, with a particular focus on the effect of BMI. Building on this, we aimed to optimize the metformin dose for treatment of type 2 diabetes in youth.

## Research Design and Methods

### Participants

Participants were recruited from the Pediatric Diabetes Clinics at the University of California, San Francisco (UCSF) from both the San Francisco and Oakland Campuses. Participants had to be between the ages of 10 to 17 years with physician diagnosed type 2 diabetes and be on immediate release metformin therapy for at least 12 weeks. Exclusion criteria included the use of extended-release metformin, the presence of pancreatic autoantibodies associated with type 1 diabetes, a diagnosis of monogenic diabetes, presence of liver disease and an estimated glomerular filtration rate (eGFR) less than 30 ml/min/1.73 m^2^. Informed consent and assent were obtained from participants. All study procedures were approved by the Institutional Review Board (IRB) at UCSF with informed consent and assent obtained from parents and the participant, respectively.

### PK sampling

Study visits took place at the Pediatric Clinical Research Center at UCSF or at participants’ homes and were conducted by trained study staff. Study procedures took place during the COVID pandemic necessitating the conduct of home visits. Participants were asked to maintain a 3-day medication log noting the exact times that they took their metformin prior to the study visit. Patients were called the day before the study visit to remind them to skip their usual morning dose of metformin on the day of the study. On the day of the study visit, height, weight and serum creatinine were measured. Participants were assumed to be at steady state at the time of the study visit and limited PK sampling was performed. Metformin levels were measured at pre-dose (trough) and a 1000 mg dose of metformin was administered. Following this, metformin levels were measured between 1–2 hours (peak) and between 3–6 hours (start of elimination) post metformin dose.

### Population PK model development

The population PK model for metformin was developed using nonlinear mixed-effects modeling approach to characterize metformin PK and sources of variability in the study population. One- and two-compartment structural models were tested, and inter-individual variability of PK parameters was assumed to follow a log-normal distribution. Participant demographics, including age, BMI, HbA1C, body surface area, and eGFR were evaluated as potential covariates using a stepwise approach with significance set at p < 0.05 for forward inclusion and p < 0.01 for backward deletion. Model fit was assessed based on the numerical and visual assessments, Including the likelihood ratio test, the goodness-of-fit plots and visual predictive checks.

### Model-based simulation

500 Monte Carlo simulations were performed to characterize the distribution of metformin exposures in the study population under different dosing strategies. 24-hour area under the concentration-time curve (AUC) were derived from simulated PK parameters and evaluated compared to those in adults under standard dosing (2,000 mg daily).

### Results

The study was conducted between June 2021 to March 2023 with a total of 50 participants completing the study. The baseline demographics of the study participants are shown in **Table 1**. 66% of the participants were male and all participants self-identified as youth of color. All the participants had a BMI Z score in the obesity range (mean 35.9 kg/m^2^ (standard deviation (SD) 6.8), range: 20 - 54) and normal kidney function (mean eGFR 114.7 mL/min/1.73 m^2^ (SD 22.6), range: 66.6 - 177.2). Most participants were on 1,000 mg of metformin twice a day. A distribution of the study doses and sampling time points are shown in **Supplementary Figures 1 and 2**. A one-compartment model with first-order absorption best described the PK data and a visual predictive check showing the model predicted values versus the actual values confirmed that our model is appropriate **(Supplementary Figure 3)**. The final PK parameter estimates are listed **Supplementary Table 1**. The raw PK data stratified by BMI categories 20-29 kg/m^2^, 30-39 kg/m^2^, and greater than 40 kg/m^2^ is shown in **Figure 1a**.

**Table 1:** The baseline demographics of the 50 study participants.

| Characteristic | Value |
| --- | --- |
| Age, years mean (SD) | 15.0 (1.6) |
| Sex, n (%) |  |
| Female | 17 (34) |
| Male | 33 (66) |
| Race and ethnicity, n (%) |  |
| American Indian/Alaska Native | 1 (2) |
| Asian | 1 (2) |
| Black | 7 (14) |
| Hispanic | 8 (16) |
| Native Hawaiian or Pacific Islander | 7 (14) |
| White | 0 (0) |
| Other/prefer not to answer | 21(42) |
| Weight, kg mean (SD) | 101.8 (22.5) |
| BMI, kg/m<sup>2</sup> mean (SD) | 35.9 (6.8) |
| BMI Z score mean (SD) | 2.3 (0.51) |
| eGFR, mL/min/1.73m<sup>2</sup> mean (SD) | 114.7 (22.6) |

SD, standard deviation; BMI, body mass index; eGFR, estimated glomerular filtration ratio

**Figure 1:**
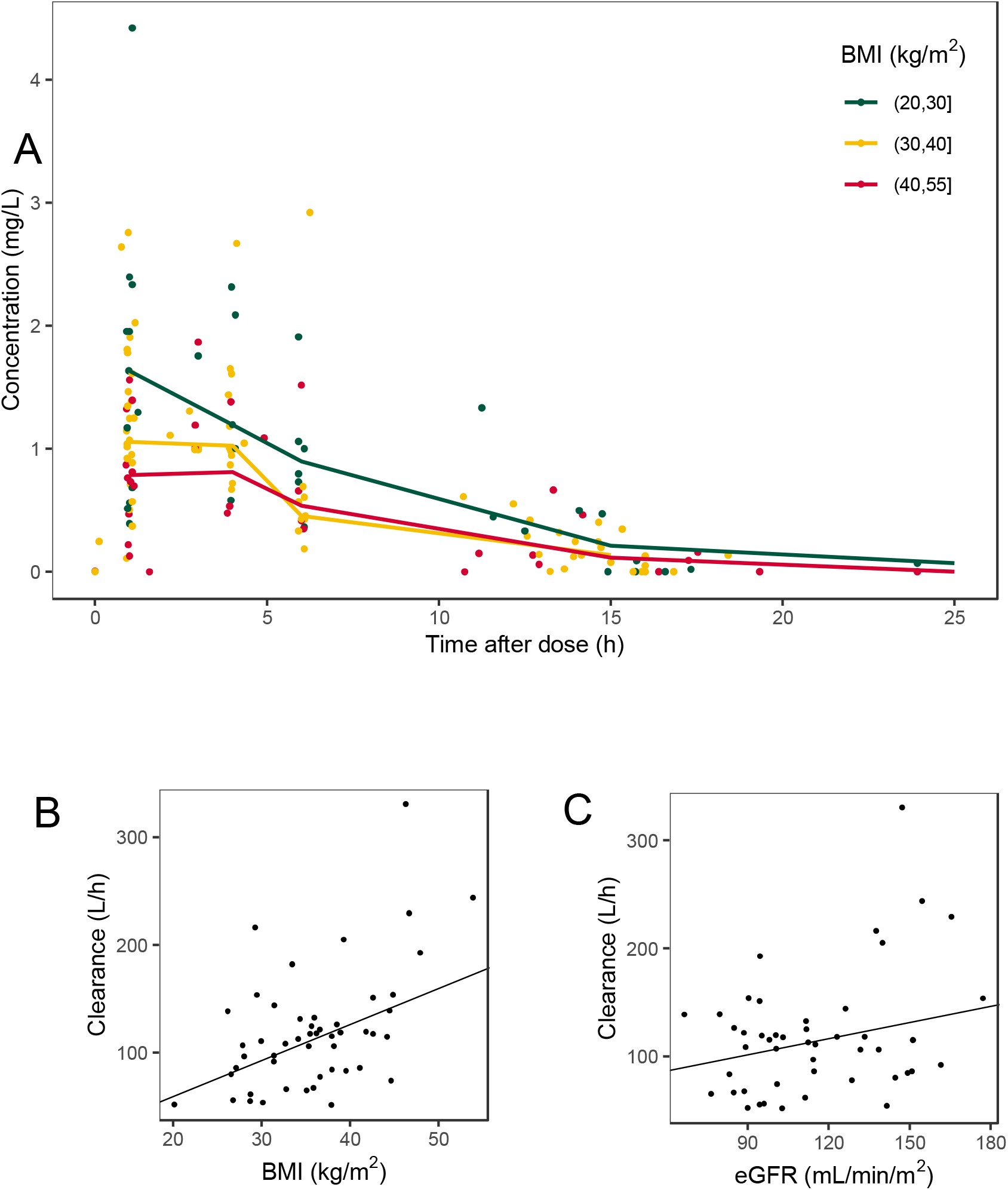
A. Raw PK data divided by BMI categories B. Relationship between clearance estimate and BMI. The line represents the model’s estimate of the effect of BMI on clearance C. Relationship between clearance and eGFR. The line represents the model’s estimate of the effect of eGFR on clearance

We evaluated the effects of potential covariates on metformin clearance and found that BMI **(Figure 1b)** and renal function **(Figure 1c)** both influence metformin clearance while age and HbA1C have no effect. Estimated clearance increased by 3% (21.4% coefficient of variation (CV)) for every 1 kg/m^2^ increase in BMI. Estimated clearance increased by 6% (53.8% CV) for every 10 mL/min/1.73 m^2^ increase in eGFR. We then compared AUC estimates from our model with those from a model developed using data from 236 adults on the same dose of 2,000 mg a day of immediate-release metformin ^6^**(Figure 2)**. Median AUC for youth receiving 2,000 mg metformin daily was lower than for adults on the same dose (18.47 mg*h/L vs. 27.49 mg*h/L). Afterwards, we used the population PK model to simulate metformin exposures in youth under different dosing strategies. Our model-based simulations showed that in youth, higher doses of at least 2,550 mg and up to 3,000 mg daily were needed to achieve exposures similar to those in adults under 2,000 mg daily **(Figure 3)**.

**Figure 2:**
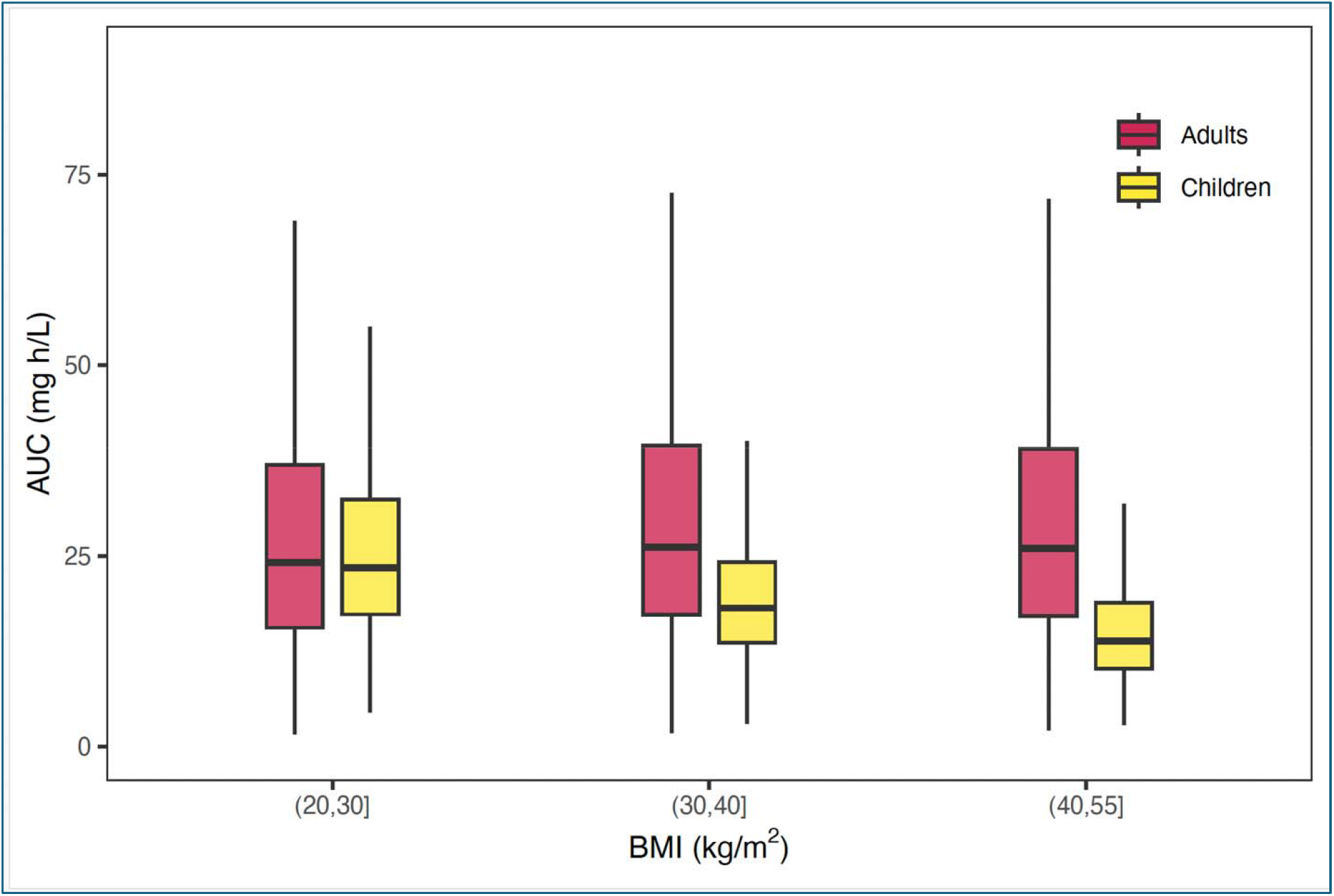
AUC estimates from our population PK model compared with those from a model developed using data from 236 adults on the same dose of 2000 mg a day of immediate-release metformin^6^

**Figure 3:**
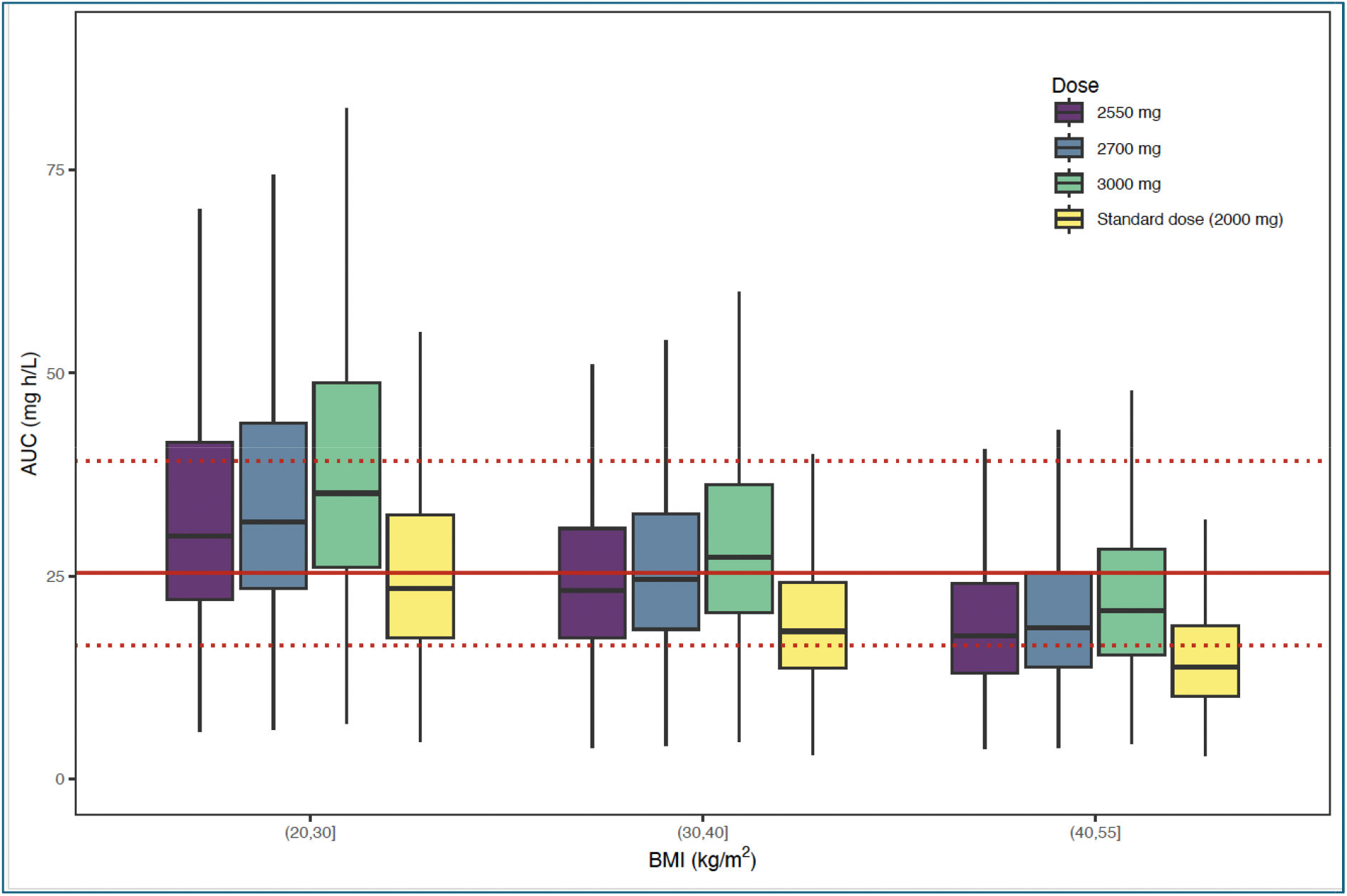
Simulated AUC for metformin doses of 2,550 mg, 2,700 mg and 3,000 mg compared to 2,000 mg. Red line represents the median adult AUC level for metformin and dotted lines represent upper and lower bounds ^1^

## Discussion

In summary, through the development of a population PK model for metformin in youth with type 2 diabetes, we show that obesity is associated with increased metformin clearance and thereby reduced metformin exposure in youth. When quantified, we see that estimated clearance increased by 3% for every unit increase in BMI. When we compared AUC estimates from our population PK model with data from adults with type 2 diabetes, we found that median AUC for youth was lower than for adults on the same 2000 mg dose across all BMI strata and in particular in the highest BMI subgroup. This suggests that youth do not achieve the same drug exposure on metformin as adults, even when given similar doses — and, in practice, they often receive lower doses. PK plays an important role in determining drug response and so this may contribute to the suboptimal metformin response observed in youth compared with adults and suggests that higher doses could enhance efficacy. Indeed, our model-based simulation indicates that doses of at least 2,550 mg, and up to 3,000 mg, are necessary to achieve metformin exposure comparable to that in adults.

Metformin was approved by the U.S. FDA for use in youth with type 2 diabetes greater than 10 years old in December 2000 based on the results of a double-blind randomized controlled trial of 82 subjects aged 10–16 years who either took a short course of metformin monotherapy or placebo pills for up to 16 weeks. The study showed that metformin significantly improved glycemic control compared to placebo with a difference of 1.1% in HbA1C levels between the metformin and placebo groups.7 However, this was a short-term study, and the dose selected was based on the most commonly used dose of 1,000 mg twice a day in adults despite the approval in adults extending to 2,550 mg a day or 850 mg three times a day. In the TODAY study, 699 youth with type 2 diabetes aged 10–17 years were randomized to one of three treatment arms after an initial run-in period to maintain everyone on metformin monotherapy: metformin up to a dose of 2,000 mg a day alone, metformin plus rosiglitazone, and metformin plus an intensive lifestyle program. One of the main findings of TODAY was that metformin monotherapy was inadequate for maintenance of glycemic control in 50% of participants after a median time to failure of ∼11 months.^8^ All the TODAY participants were overweight or had obesity and so it may be possible that participants did not have achieve adequate metformin drug exposure contributing to non-response.

Physiological changes associated with obesity can significantly impact drug PK and, consequently, pharmacodynamics, potentially necessitating dose adjustments to achieve optimal therapeutic effects. However, there is limited labeling information available for metformin in youth with obesity and type 2 diabetes. Our model-estimated metformin PK parameters are consistent with reports from previous small studies in youth which have shown that body weight and estimated glomerular filtration rate are significant covariates that affect metformin clearance and AUC parameters.^9,10^ To our knowledge, our study represents the first PK study of metformin in youth with type 2 diabetes. A recent study applied and scaled a physiologically based PK model of metformin to data digitized from three metformin PK studies with data from children both with and without obesity. They found that predicted metformin clearance was higher and AUC lower in children with obesity, but the corresponding values were comparable to findings in adults.^11^ Several differences exist between this study and ours. This study was conducted using a digitized dataset of children without type 2 diabetes and the range of overweight and obesity was lower than observed in our study. Additionally, the sample size was smaller and therefore available data for model development was sparser potentially leading to differences in findings.

Metformin is a drug that is primarily eliminated through renal tubular excretion. A potential mechanism for increased clearance observed in children with obesity could be related to greater kidney volume and glomerular filtration rates which have been broadly documented in obese individuals.^12,13^ FDA labels for pediatric indications are typically at doses similar to or lower than approved in adults. However, children are not young adults and in particular type 2 diabetes in youth predominantly occurs after the onset of puberty with youth tending to have a greater degree of adiposity than their adult counterparts. Given these findings, our results may help guide dosing strategies for other renally cleared drugs in youth with type 2 diabetes, as well as medications used in children with obesity across various disease states, especially in light of the increasing prevalence of obesity.

Metformin remains one of the most widely prescribed diabetes drugs in the US and worldwide. While newer medications are becoming increasingly available, they are still difficult to access and expensive. Therefore, efficacious use of existing drugs is important to optimize care particularly in resource limited settings. Strengths of our study include that it involved a large and diverse sample of 50 youth with type 2 diabetes with rigorous PK sampling methodology and model development. Limitations include that extended-release metformin formulations were not tested and that we were not able to comment on differences in PK based on sex or self-reported race and ethnicity with our sample size.

In conclusion, our findings demonstrate that metformin PK vary by BMI status in youth with type 2 diabetes, with obesity resulting in reduced drug exposure levels. The next important step is to determine whether a higher metformin dose can be tolerated and elicits improved glycemic response in youth. To address this, we are currently conducting a pilot double-blind randomized controlled trial, Precision Dosing of Metformin in Youth with Type 2 Diabetes (PRECISE_T2D) (NCT06120881) to evaluate a higher dose of metformin (2,700 mg a day) compared with standard dosing (2,000 mg a day) in 20 youth with obesity and type 2 diabetes over a period of three months.

## Data Availability

All data produced in the present study are available upon reasonable request to the authors

