## Supplemental Files for "Obesity is associated with increased clearance and reduced drug exposure to metformin in youth with type 2 diabetes"

**Supplementary Table 1: Pharmacokinetic parameter estimates of the metformin population pharmacokinetic model**

| **Parameter** | **Estimate (% RSE)** |
| --- | --- |
| **Clearance (L/h)** | 110 (8.8) |
| **Volume of distribution (L)** | 461 (10.7) |
| **Absorption rate constant, k_a_ (/h)** | 0.76 (11.8) |
| **% Increase in Clearance per 1 kg/m^2^ increase in BMI** | 3 (45.4) * |
| **% Increase in Clearance per 10 mL/min/1.73 m^2^ increase in** **eGFR** | 6 (53.5) * |

*P < 0.05

RSE, relative standard error; BMI, body mass index; eGFR, estimated glomerular filtration ratio

**Supplementary Figure 1: Distribution of metformin doses taken by study participants**

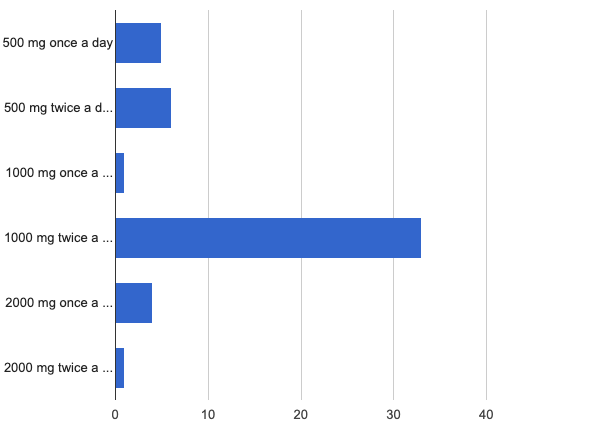

Metformin dose

Number of measurements

**Supplementary Figure 2: Distribution of pharmacokinetic sampling time points**

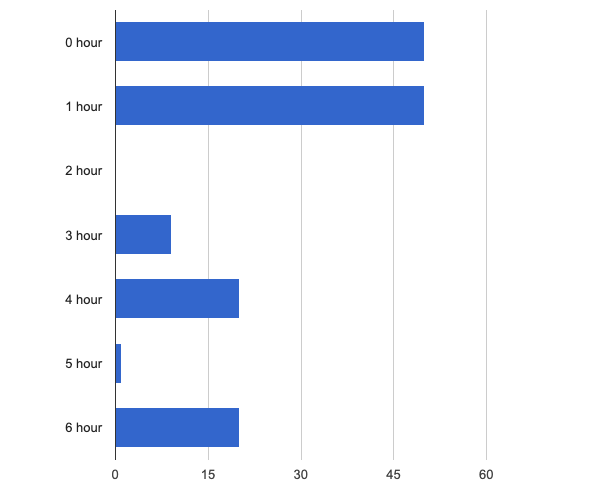

Time point

Number of measurements

**Supplementary Figure 3: Visual predictive check showing the model predicted values versus the actual values**

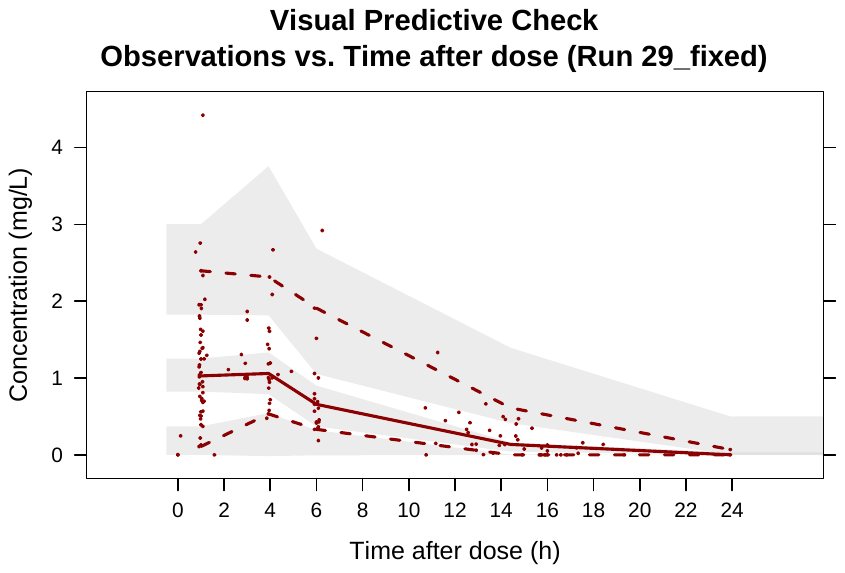
